# Resting-state EEG and machine learning to investigate cortical connectivity as a biomarker in chronic mTBI

**DOI:** 10.1101/2025.10.28.25338644

**Authors:** William J. Marshall, Amy N. Conner, Alexandra P. Key, Tonia S. Rex

## Abstract

Mild traumatic brain injury (mTBI) is a heterogeneous condition with long-term sequelae, yet diagnosis in the chronic stage remains limited by reliance on acute criteria and subjective reports. Objective biomarkers are needed, as current blood-based markers show diagnostic value primarily in the acute and subacute phases. Resting-state EEG (RS-EEG) can capture large-scale network disruptions through functional connectivity (FC) and microstate analysis, but its role in chronic mTBI is unclear. We tested whether RS-EEG features distinguish chronic mTBI from controls and predict symptom burden. This observational case-control study included 44 participants (18 chronic mTBI, 26 controls). Source-reconstructed EEG was analyzed for spectral power, microstate metrics, and FC using the Multivariate Interaction Measure (MIM). Elastic Net and XGBoost models classified injury status and predicted symptom severity, with feature robustness evaluated across full and reduced electrode montages. Participants with mTBI showed no group differences in spectral power or microstate metrics but demonstrated significantly elevated FC across theta, beta, gamma, and broadband frequencies. Connectivity increases were stable across reduced montages and persisted up to eight years post-injury. Classification models using MIM achieved AUCs of 0.79–0.89 for injury status and 0.82–0.87 for symptom severity, outperforming demographic models. Resting-state EEG FC provides a sensitive biomarker of chronic mTBI, distinguishing cases from controls and correlating with symptom severity. The persistence of network alterations years after injury suggests lasting changes in brain activity associated with chronic symptom burden. These findings support the use of RS-EEG–derived FC as a noninvasive and scalable biomarker of chronic mTBI.

## INTRODUCTION

Mild traumatic brain injury (mTBI) is a heterogeneous neurological condition with potential for persistent symptoms and neurophysiological disruption.^1^ Yet, diagnosis in the chronic stage remains challenging. Acute measures such as the Glasgow Coma Scale (GCS) are not administered, and neuroimaging is often not obtained, leaving clinicians to rely on symptom reports that may not reflect underlying brain dysfunction. Up to 35–53% of individuals remain symptomatic more than 5–10 years post-injury,^2,3^ emphasizing the need for objective biomarkers to detect and monitor chronic impairment.

The recently proposed clinical-biomarker-imaging modifiers (CBI-M) framework highlights the importance of functional biomarkers to complement symptom-based criteria to enhance classification accuracy.^4^ While blood-based biomarkers such as glial fibrillary acidic protein (GFAP) and ubiquitin C-terminal hydrolase L1 (UCH-L1) show diagnostic utility in the acute (within 24 hrs) and subacute (within 3 months) phases of mTBI,^5^ their role in identifying or monitoring long-term dysfunction (> 6 months – 1 year) remains underexplored. Neurophysiological measures, particularly resting-state EEG (RS-EEG), offer a scalable and noninvasive alternative for probing persistent network dysfunction.

RS-EEG provides a window into large-scale brain dynamics. Functional connectivity (FC) analyses quantify the efficiency of communication between brain regions, while microstate analysis characterizes rapid transitions between global brain states.^6,7^ Both FC and microstates show alterations across neurological and psychiatric disorders, with changes manifesting as either increases or decreases depending on the condition.^8–12^ However, their utility in chronic mTBI, particularly in individuals with diverse injury mechanisms and extended post-injury durations, remains unclear. mTBI is known to disrupt distributed cortical hubs involved in attention, sensory integration, and executive function,^13^ and our prior work in this cohort identified atypical auditory and visual evoked responses years after injury,^14,15^ suggesting that sensory processing deficits may reflect broader network dysfunction.

To address this gap, we hypothesized that individuals with chronic mTBI would show altered RS-EEG connectivity and microstate dynamics, reflecting persistent network disruption. Because scalp EEG signals can be spatially ambiguous, we applied source reconstruction to enhance anatomical specificity, enabling biologically interpretable network-level analyses. We further tested whether these features could classify injury status, predict symptom burden, and remain robust under reduced electrode montages, supporting their potential as clinically scalable biomarkers.

## METHODS

### Participants

This observational case-control diagnostic study enrolled adults aged 18-72 with either a history of mild traumatic brain injury (mTBI; GCS 13-15) or no history of head injury (controls).

Exclusion criteria included severe TBI (GCS <13), eye or ear disease (e.g., Meniere’s disease, glaucoma), metallic implants incompatible with neuroimaging, pregnancy, or age <18. All participants had normal hearing thresholds and 20/20 best-corrected visual acuity. Corrective lenses were permitted.

A total of 65 individuals were recruited, with 28 participants with mTBI (mean age 35, SD = 13.08, range = 19-72, 13 female) and 34 controls (mean age 39, SD = 11.34, range = 23-71, 25 female) included after exclusions. Out of 62 included participants, resting-state EEGs were completed for 23 participants with mTBI and 29 controls. mTBI history was confirmed via medical records or structured history, including injury mechanism and treatment. Mechanisms included motor vehicle accidents (*n* = 9), falls (*n* = 7), non-violent blunt trauma (*n* = 3), and sports injuries (*n* = 4). The average time since last mTBI was 7.25 ± 6.71 years.

### Procedure

Participants completed the Neurobehavioral Symptom Inventory (NSI), a validated 22-item self-report measure of post-concussive symptoms.^16^ Resting-state EEG (RS-EEG) was recorded in a quiet, sound-treated room. Data were collected with a 128-channel hydrocel net (Electrical Geodesics, Inc., Eugene, OR) at 1000 Hz, referenced to Cz. Electrode impedances were kept <50 kΩ. Recordings included 3 minutes eyes-open (EO) followed by 3 minutes eyes-closed (EC). The fixed order minimized drowsiness and artifact variability. During EO, participants fixated on a central cross to reduce eye movements. Condition transitions were signaled by a 500 Hz auditory beep, with event markers inserted into the EEG file for alignment. This was part of a larger study, components of which have been published elsewhere.^14,15^

### EEG Preprocessing and Source Reconstruction

#### Preprocessing

Data were preprocessed in MATLAB (MathWorks, Natick, MA) using EEGLAB^17^ and a customized version of the Maryland Analysis of Developmental EEG (MADE) pipeline.^18^ As part of the MADE pipeline, signals were bandpass filtered at 0.1-50 Hz using the FIRfilt plugin,^19^ then segmented into 2000-ms epochs. Bad channels were identified and interpolated, and epochs with >10% interpolated electrodes or >±100 μV amplitude were rejected. The Fully Automated Statistical Thresholding for EEG artifact Rejection (FASTER) algorithm aided channel-level artifact detection,^20^ and artifact-related independent components were identified using ADJUST.^21^ Eyes-closed recordings were used for primary analyses due to lower artifact susceptibility.^22^

#### Source Reconstruction

Source reconstruction was performed using ROIConnect^23^ (EEGLAB plugin) with the Montreal Neurological Institute (MNI) Boundary Element Model^24^ and the Colin27 template.^25^ Cortical activity was estimated via exact low-resolution electromagnetic tomography (eLORETA)^26^ and parcellated into Desikan-Killiany^27^ atlas regions to extract biologically interpretable ROI time series. All subsequent analyses of spectral power, microstates, and functional connectivity were conducted using these source-reconstructed ROI time series.

### EEG Analyses

#### Power Spectra Analysis

Power spectral density (PSD) was computed to assess frequency-specific neural oscillations, given prior reports of spectral alterations in neurotrauma.^28,29^ Data were analyzed in EEGLAB with 20,000 permutations and False Discovery Rate (FDR) correction (p < 0.05).^30^ The FOOOF toolbox was used to separate narrowband oscillations from the aperiodic 1/f component.^31^ Power was examined across delta (1–4 Hz), theta (4–8 Hz), alpha (8–12 Hz), beta (13–30 Hz), and gamma (30–50 Hz) bands.

#### Microstate Analysis

Microstate analysis across 1-50 Hz was conducted in MicrostateLab (EEGLAB plugin).^32^ Global Field Power peaks were clustered (k = 4–7) using k-means, with optimal solutions selected via meta-criteria in Cartool.^33^ Group-averaged maps were matched to canonical templates based on the highest correlation, and temporal dynamics (duration, coverage, occurrence) were derived by back-fitting. Group differences were assessed using Topographic Analysis of Variance (TANOVA) in RAGU with 100,000 permutations and FDR correction.^34^

#### Functional Connectivity (MIM)

Functional connectivity was computed using the Multivariate Interaction Measure (MIM), which reduces the impact of volume conduction by focusing on non-instantaneous interactions in the frequency domain. Connectivity was estimated for the broadband range (1–50 Hz) and standard frequency bands (delta [1-4 Hz], theta [4-8 Hz], alpha [8-12 Hz], beta [13-30 Hz], gamma [30-50 Hz]). Outliers were capped at 3×IQR to reduce undue influence. Group differences were tested with type III ANCOVA controlling for age and sex. Because Levene’s test indicated heteroscedasticity, robust standard errors were applied.^35^ Effect sizes were estimated with bootstrapped partial η² and 95% confidence intervals.

#### Reduced ROI Analysis

To evaluate clinical scalability, we repeated FC analyses using reduced ROI subsets designed to approximate sparse EEG montages. These included occipital (visual) (1), superior temporal (auditory) (2), and frontoparietal regions (3) (Figure 1). Results were FDR-corrected across frequency bands and montage conditions.

**Figure 1.**
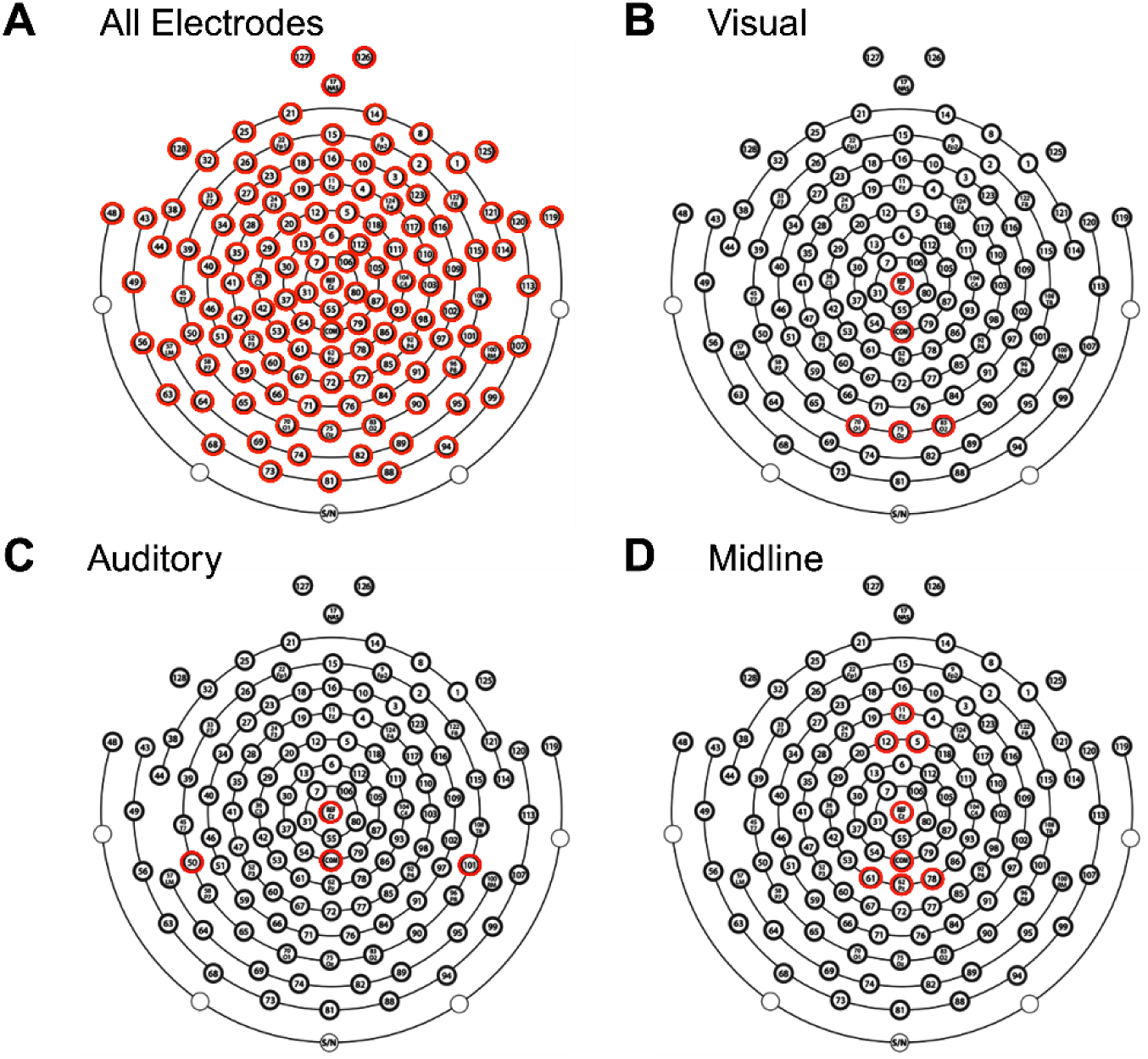
Electrode Selection for EEG Analysis. Topographical layout of the EEG cap showing electrodes included in four analyses. (A) All electrodes, (B) visual subset, (C) auditory subset, and (D) midline subset. Highlighted electrodes (red) indicate those used in each respective analysis, including the reference (Cz) and common ground electrode (COM).

#### Classifier Analysis

To evaluate whether FC features distinguished chronic mTBI from controls, we applied three complementary classification methods: (1) threshold-based cutoffs, which use a single optimal decision boundary (Youden’s index) to maximize sensitivity and specificity;^36^ (2) Elastic Net logistic regression, a linear model that combines L1/L2 regularization to handle correlated predictors and reduce overfitting;^37^ and (3) XGBoost gradient-boosted decision trees, a nonlinear ensemble method capable of modeling higher-order feature interactions.^38^ This combination allowed comparison of simple interpretable models versus more flexible machine learning approaches. MIM values were z-scored for Elastic Net but left unscaled for XGBoost. Hyperparameters were tuned with 5-fold cross-validation. Classification performance was quantified with ROC receiver operating characteristic (ROC) curves and area under the curve (AUC, 95% CIs from 20,000 bootstraps). Model significance was assessed with 20,000 permutation tests of shuffled labels. Models were implemented in MATLAB (Elastic Net) and Python (XGBoost with scikit-learn and xgboost).

#### Symptom Severity Prediction Analysis

Associations between FC and post-concussive symptoms were examined using NSI total score. Linear regression tested whether MIM values predicted total NSI scores (controlling for age and sex). For secondary analyses, participants were dichotomized into no-mild (NSI total score <25) vs. moderate-severe (NSI total score ≥25) symptom groups, and classification was performed using Elastic Net and XGBoost. Models were tuned with 5-fold cross-validation, and performance was quantified with ROC AUCs (95% CIs via 20,000 bootstraps). Statistical significance was assessed with 20,000 label-permutation tests.

### Standard Protocol Approvals, Registrations, and Patient Consents

The study was approved by the Vanderbilt University Medical Center Institutional Review Board (IRB #171061). All participants provided written informed consent.

### Reporting Guidelines

This study was conducted in accordance with the STROBE (Strengthening the Reporting of Observational Studies in Epidemiology) and STARD (Standards for Reporting Diagnostic Accuracy Studies) guidelines. Completed checklists are available upon request.

### Data Availability

Anonymized data not published within this article will be made available on request from qualified investigators.

## RESULTS

After exclusions for incomplete recordings (*n* = 5, 2 controls) and excessive artifacts (*n* = 3, 1 control), 44 participants were included in the final analysis. The control group (*n* = 26) had a mean age of 39.6 years (SD = 11.5) and was 73% female, while the mild TBI group (*n* = 18) had a mean age of 32.0 years (SD = 11.4) and was 50% female. Age distributions were compared between included and excluded participants. For mTBI participants, Shapiro–Wilk indicated non-normality in the included group (p = 0.003), so a Mann-Whitney U test was used. Excluded cases were significantly older (U = 53.0, p = 0.041). For controls, both groups were approximately normal (p > 0.07), and Welch’s t-test showed no difference (t = 0.88, p = 0.39). Thus, the final analysis sample contained younger mTBI participants relative to excluded cases, while control demographics were preserved. To account for this potential confound, age was included as a covariate in all analyses.

Among mTBI participants, the average time since their last mTBI was 7.7 years (SD = 7.0), consistent with chronic-phase recovery. Injury mechanisms are detailed in Supplementary Table 1. There were no significant differences in injury time or mechanism between excluded and included participants with mTBI (p > 0.05).

After preprocessing, average data retention was lower in cases than controls (mTBI: mean = 0.68, SD = 0.32; Control: mean = 0.92, SD = 0.12; Mann–Whitney U test, p = 0.009). For eyes-open EEG, retention did not differ between groups (mTBI: mean = 0.78, SD = 0.28; Control: mean = 0.86, SD = 0.17; Mann–Whitney U test, p = 0.424). Retention was lower in mTBI participants during eyes-closed recordings, likely reflecting increased artifact, but all participants retained adequate data for valid analysis. Moreover, the consistency of eyes-open retention across groups suggests that the observed group differences are not attributable to general data quality disparities.

### Resting-state spectral power is preserved after chronic mTBI

No significant group differences were observed in eyes-closed resting-state power spectra across the full 0–50 Hz range (*p* > 0.05; Figure 2A), indicating that overall spectral content remained comparable between mTBI and control participants. This pattern held after removing the aperiodic background using FOOOF, isolating true oscillatory components. Group comparisons of the periodic spectra also revealed no significant differences (*p* > 0.05; Figure 2B), suggesting that mTBI does not produce broad alterations in canonical frequency bands during rest.

**Figure 2.**
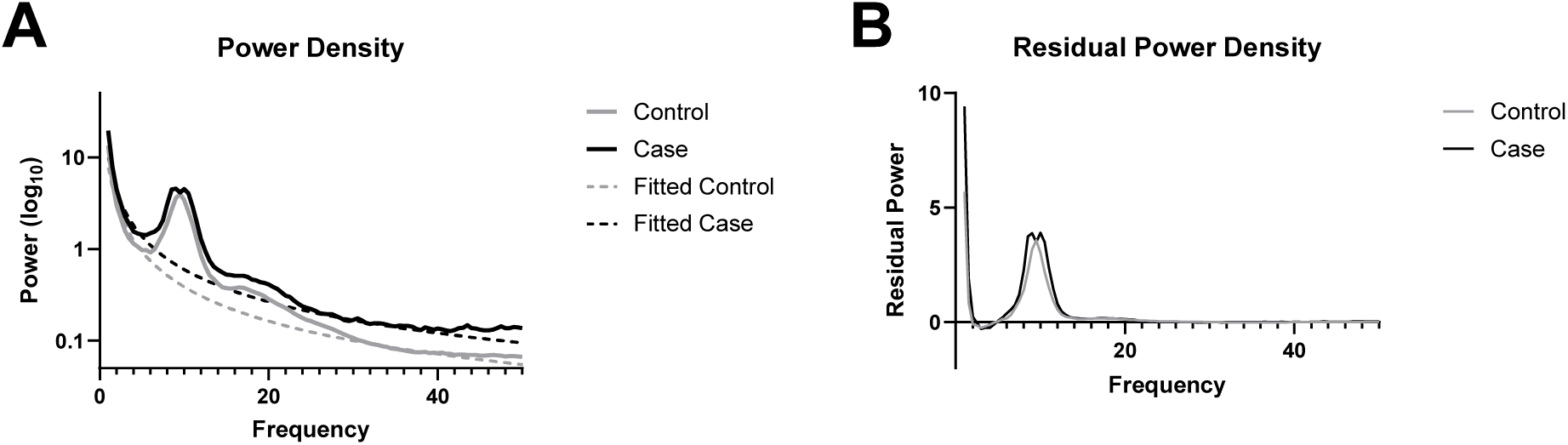
Power Spectral Density. A: Power spectra with an aperiodic fit overlay. B: Periodic residual from the removal of the aperiodic fit from raw power spectra

### Microstate topographies and temporal dynamics are unaltered after chronic mTBI

Microstate topographies, reflecting canonical spatial configurations of scalp potential fields, were statistically indistinguishable between the mTBI and control groups across 5 optimal microstates, as assessed using TANOVA (Figure 3A-B). This indicates that the overall spatial organization of microstates was preserved in individuals with chronic mTBI. Further, the temporal dynamics of the backfitted microstate sequences, including duration, occurrence, coverage, global explained variance, and transition probabilities, also showed no significant group differences (Figure 3C-F), suggesting preserved microstate temporal dynamics in individuals with chronic mTBI.

**Figure 3.**
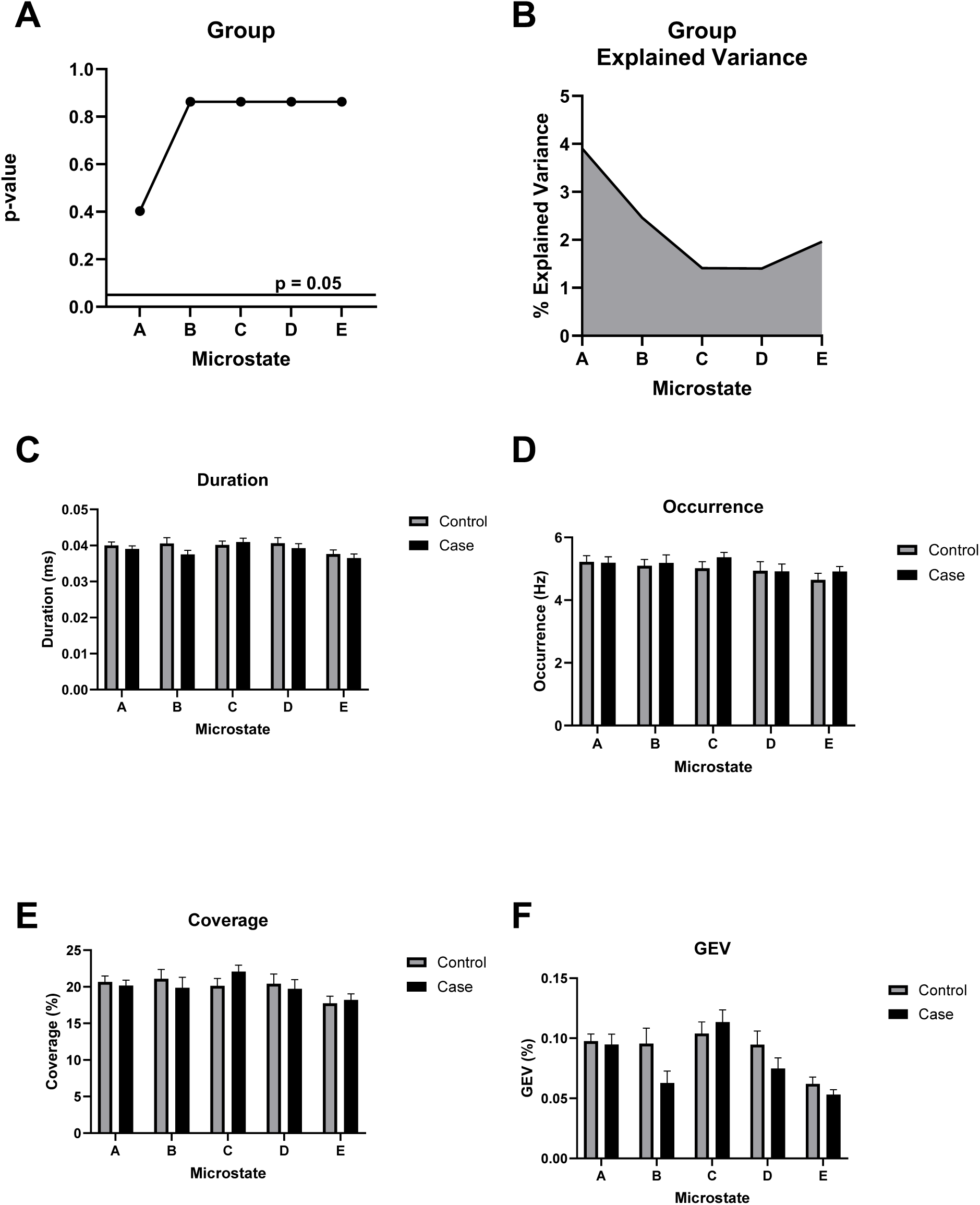
Microstate Topographical Analysis of Variance (TANOVA). A: *p*-value of the topographical difference based on group (mild TBI/Control) across all microstates. All *p*-values use FDR correction. B: Cumulative explained variance (CEV) based on group. C: Duration (in milliseconds) reflects the average length of each microstate. D: Occurrence (in Hz) represents the frequency of each microstate per second. E: Coverage denotes the proportion of total time occupied by each microstate. F: Global Explained Variance (GEV) quantifies the proportion of EEG variance explained by each microstate. No significant group differences were observed across any microstate metric (A-F).

### Chronic mTBI is associated with widespread cortical hyperconnectivity

The mild TBI group exhibited significantly higher functional connectivity compared to controls across all frequencies and within theta, beta, and gamma bands (p < 0.05), with no group differences observed in the delta or alpha bands (Figure 4A). In frequency bands showing significant effects, group membership (i.e., mTBI or control) accounted for an average of 21% of the variance (Figure 4B). These group differences remained robust when using reduced ROI subsets approximating standard EEG configurations, including a visual montage based on visual evoked potentials (VEP), an auditory montage based on auditory evoked potentials (AEP), and a midline- only montage (Figure 4C). Across these subsets, group explained an average of 23% of the variance in connectivity, again reflecting increased connectivity in the mTBI group relative to controls (Figure 4D).

**Figure 4.**
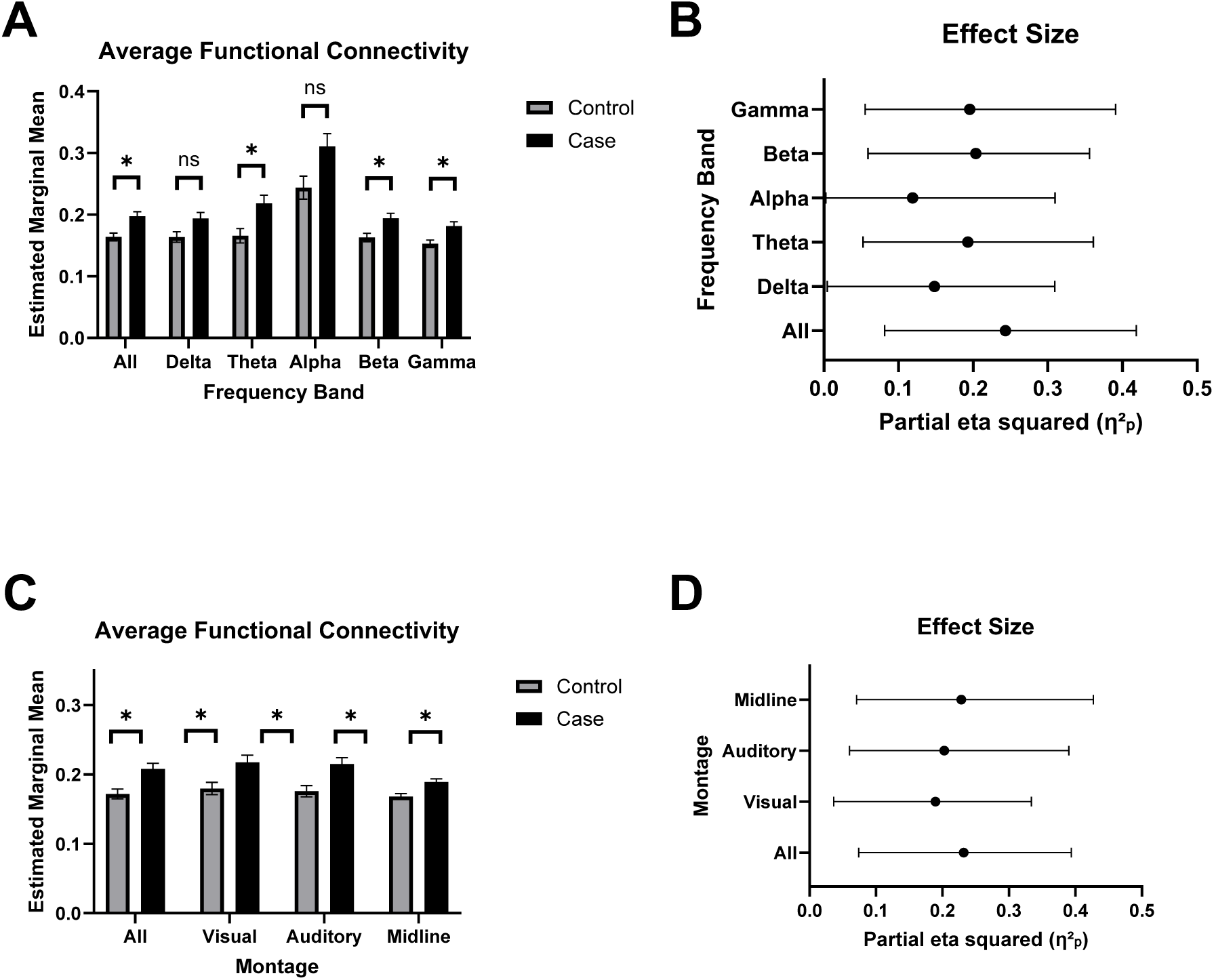
Increase in Estimated Functional Connectivity. A: Estimated marginal mean MIM (Multivariate Interaction Measure) across frequency bands: delta (1–3 Hz), theta (4–7 Hz), alpha (8–12 Hz), beta (13–30 Hz), and gamma (30–45 Hz). Bars represent mean ± SEM for each group. Asterisks indicate FDR-corrected statistical significance between groups (p < 0.05 = *, p < 0.01 = **, p < 0.001 = ***). B: Effect sizes for each frequency band are reported as partial eta squared (η²ₚ), indicating the proportion of variance accounted for by MIM. Bars represent mean ± SEM for each group. C: Estimated marginal mean MIM across subregions to simulate alternative EEG montages. D: Effect sizes for each montage are reported as partial eta squared (η²ₚ), indicating the proportion of variance accounted for by MIM.

### Machine Learning captures robust differences in functional connectivity after chronic mTBI

We evaluated the utility of functional connectivity (MIM) for classifying chronic mTBI from control participants using three classification approaches: a threshold-based classifier, Elastic Net regression, and XGBoost. Using the raw MIM values, the optimal threshold (0.174) yielded an AUC of 0.793 (95% CI: 0.615-0.940) (Figure 5A). The Elastic Net model (α = 0.03) produced a similar AUC of 0.790 (95% CI: 0.524 – 1.000) (Figure 5C). Both the threshold-based and Elastic Net models are acceptable and close to the 0.8 threshold commonly interpreted as excellent classification performance.^39^ The XGBoost model achieved the highest accuracy with an AUC of 0.887 (95% CI: 0.785-0.943) (Figure 5E). SHAP analysis indicated that higher MIM values were strongly associated with increased likelihood of mTBI classification (Figure 5F), reinforcing the discriminative power of functional connectivity, even in more complex models. All permutation tests yielded p-values less than 0.05, confirming that the observed AUCs were unlikely to occur by chance (Figure S2A-C).

**Figure 5.**
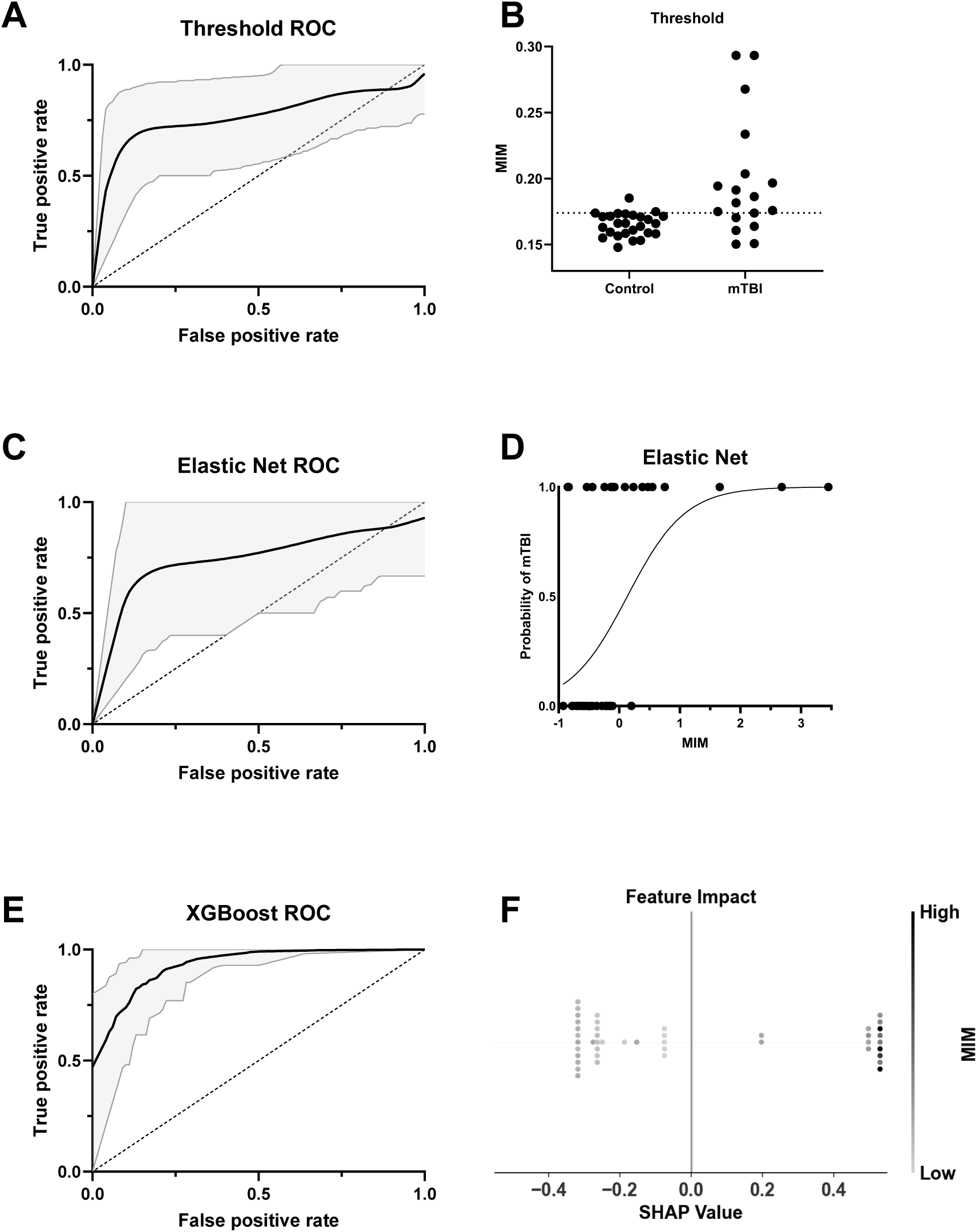
Classifier performance and interpretability for group prediction based on MIM. A: ROC curve for a threshold-based classifier using the optimal Youden’s J statistic from z-scored MIM. The black line indicates the mean ROC curve across 20,000 bootstraps, with 95% confidence interval shaded in gray. B: Distribution of normalized MIM values across Control and mTBI groups. The dashed line indicates the optimal threshold used in A. C: ROC curve for an Elastic Net classifier using MIM as the predictor. The solid line shows the average ROC across bootstraps, with 95% CI shaded. D: Probability curve for Elastic Net classifier output as a function of MIM. The logistic decision function exhibits a higher probability of mTBI with increasing MIM. E: ROC curve for an XGBoost classifier using MIM as the predictor. F: SHAP summary plot from the XGBoost model. Each dot represents an individual’s SHAP value, indicating feature impact on model output, with color based on the strength of z-scored MIM.

Model performance varied when age and sex covariates were included. Elastic Net performance decreased slightly, with an AUC of 0.765 (95% CI: 0.517 – 0.967) (Figure S3A), possibly due to overfitting or noise introduced by additional predictors in the context of a modest sample size. In contrast, XGBoost performance improved, reaching an AUC of 0.901 (95% CI: 0.844 – 0.944) (Figure S3C), suggesting better accommodation of additional covariates. Permutation tests again confirmed model significance (p < 0.05) (Figure S3B and D). When MIM was removed from the models, performance declined substantially: the Elastic Net dropped to an AUC of 0.641 (95% CI: 0.438–0.867) and XGBoost to 0.835 (95% CI: 0.701–0.912; Figure S4A, C). Despite this, models remained statistically significant (p < 0.05). Collectively, these results highlight MIM’s strong predictive contribution across models.

### Cortical hyperconnectivity after chronic mTBI reflects neurobehavioral symptom severity

MIM significantly predicted total symptom burden as measured by the Neurobehavioral Symptom Inventory (NSI) total score in a linear regression model (Figure 6A). Symptoms were distributed across emotional, cognitive, sensory, and vestibular domains (Figure 6B). MIM also effectively distinguished mTBI patients with moderate to severe symptoms (NSI total score ≥ 25) from those with mild or no symptoms (NSI total score < 25) when used as a predictor in both Elastic Net logistic regression (AUC = 0.824 [95% CI: 0.33-1]) and XGBoost classifier models (AUC = 0.865 [95% CI: 0.80-0.95]; Figure 6C and 6E). Permutation testing confirmed that the observed model performance was statistically significant (p < 0.05; Figure 6D and 6).

**Figure 6.**
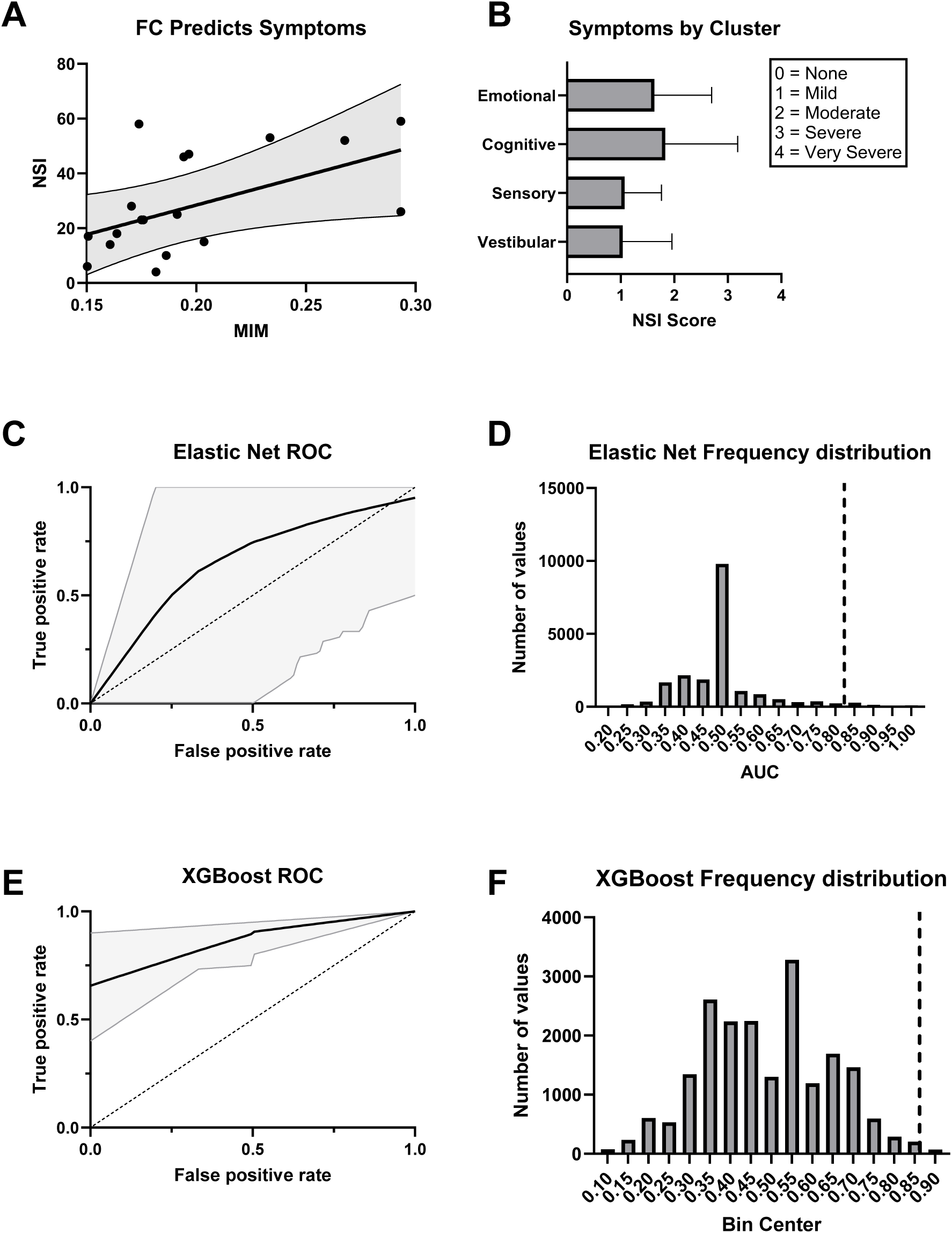
Functional Connectivity explains Neurobehavioral Symptoms. A: Higher functional connectivity (MIM) predicts greater symptom severity as measured by the Neurobehavioral Symptom Inventory (NSI). The regression line is shown with 95% confidence intervals. B: Average NSI scores across symptom clusters. Scores are rated on a 0–4 scale, with 0 = None, 1 = Mild, 2 = Moderate, 3 = Severe, and 4 = Very Severe. Error bars represent ±1 SEM. C: ROC curve for an Elastic Net logistic regression classifier trained on MIM. The black line indicates the mean ROC curve across 20,000 bootstraps, with 95% confidence interval shaded in gray. B: Histogram displaying the frequency of AUC values under the null hypothesis for the Elastic Net model. The dashed vertical line indicates the observed AUC for the true model in each case. The observed AUC had statistically significant model performance above chance (p < 0.05). C: ROC curve for an XGBoost classifier trained on MIM. The black line indicates the mean ROC curve across 20,000 bootstraps, with 95% confidence interval shaded in gray. D: Histogram displaying the frequency of AUC values under the null hypothesis for the XGBoost model. The dashed vertical line indicates the observed AUC for the true model in each case. The observed AUC had statistically significant model performance above chance (p < 0.05).

Model performance declined when demographic covariates were added. The Elastic Net model, including MIM, age, and sex, yielded an AUC of 0.661 (95% CI: 0–1) and did not reach statistical significance (p = 0.125; Figure S5A–B). A model using only age and sex performed worse than chance (AUC = 0.421, 95% CI: 0–0.583; p = 0.698; Figure S6A–B). In contrast, the XGBoost model that included MIM, age, and sex maintained strong performance (AUC = 0.874, 95% CI: 0.733–1; p < 0.05; Figure S5C–D), while the XGBoost model trained solely on demographics showed reduced accuracy (AUC = 0.732, 95% CI: 0.625–0.833) and was not statistically significant (p = 0.147; Figure S6C–D). These results demonstrate that including MIM as a predictor significantly improved classification of symptom severity, reinforcing its potential utility as a functional biomarker in chronic mTBI.

## DISCUSSION

### Overview and key findings

This study investigated whether RS-EEG features, including spectral power, microstate metrics, and functional connectivity, can differentiate individuals with chronic mTBI from healthy controls. Using source-reconstructed RS-EEG, we first assessed group-level differences. Subsequently, we utilized machine learning models to evaluate the capability of FC metrics to classify injury status and predict symptom severity. Although spectral power and microstate metrics did not reveal significant differences between groups, functional connectivity measured via MIM demonstrated robust and widespread hyperconnectivity in the mTBI group. These were identified up to nearly eight years post-injury and significantly predicted symptom severity and injury status, highlighting EEG-derived connectivity as an ongoing challenge for mTBI patients that may contribute to chronic symptomology.

### Spectral power and microstate metrics lack sensitivity in chronic mTBI

No group differences in spectral power were observed, consistent with prior reports in both the subacute (<3 months) and chronic (>10 years) phases.^29,40^ Although group means were unchanged, spectral measures have been linked to symptoms, including associations with memory, injury severity, and psychological problems.^29,40^ Other studies, however, have identified abnormalities, such as increased theta and reduced beta power in the acute phase, altered frontal alpha/beta asymmetry persisting beyond 9 months, and reduced beta with elevated delta/gamma activity in adolescents up to 12 months post-injury.^41–43^ These discrepancies, together with the fact that spectral power primarily reflects local rather than network-level activity,^13^ suggest limited sensitivity to the distributed network disruptions typical of mTBI. Accordingly, spectral power appears to have low value for screening or longitudinal monitoring, and measures of functional connectivity may provide greater specificity and clinical relevance.

Similarly, canonical EEG microstate metrics showed no group differences. Spatial configurations and temporal parameters (duration, occurrence, coverage) were preserved in chronic mTBI. Effects have been reported when analyses were stratified by symptom severity, including shorter microstate durations and reduced alpha/theta activation in patients with moderate-to-severe neuropsychological impairments.^44^ However, because such effects are not evident at the group level, canonical microstates also appear limited as stand-alone biomarkers. By contrast, network-based microstates derived from MEG, defined from whole-brain connectivity rather than scalp voltage maps, have shown higher classification accuracy.^45^ While this approach may better capture large-scale network disruptions, MEG’s cost and limited availability constrain its clinical feasibility.

### Persistent functional hyperconnectivity in chronic mTBI

Our MIM-based functional connectivity analysis revealed significant and widespread increases in broadband, theta, beta, and gamma bands in the mTBI group. This pattern is strikingly similar to Wu et al.,^46^ who also reported increased connectivity in theta, beta, and gamma, but not alpha or delta, in professional boxers with a history of repetitive mTBI. The alignment across studies may reflect shared neural signatures of chronic-stage injury, as both examined participants more than a year post-injury and with likely cumulative exposure. One distinction is that Wu et al. observed hemisphere-specific effects, with stronger FC in the right hemisphere, potentially reflecting asymmetric head impacts in boxing. Our cohort’s more varied injury mechanisms may have produced a more bilateral pattern.

In contrast, studies of patients in the acute-to-subacute phase have sometimes reported reduced connectivity across multiple frequency bands.^45^ Such decreases are less common than early hyperconnectivity, which is often observed soon after injury and is thought to reflect compensatory recruitment of alternative pathways.^13^ Methodological differences may also play a role. Dynamic MEG-based analyses may be more sensitive to localized hub or long-range disruptions, whereas static EEG-based approaches capture broader global patterns. Stage of injury, analytic method, and sample characteristics likely explain why reductions are more apparent acutely, while chronic-phase data, including the present results, tend to show hyperconnectivity.

The compensatory hyperconnectivity model suggests that early increases in connectivity support recovery, but persistence into the chronic phase may signal inefficient signaling, maladaptive reorganization, or ongoing dysregulation.^13^ The absence of alpha and delta effects in chronic cohorts may further indicate that these bands are more sensitive to acute-stage disruptions, while later variability reflects divergent recovery trajectories and injury profiles.

Source-level EEG connectivity metrics such as MIM align more closely with anatomical networks than traditional sensor-level approaches,^47^ reinforcing the validity of the present findings. Future multimodal work combining EEG with diffusion MRI will be critical to determine whether chronic hyperconnectivity reflects sustained compensation, unresolved structural injury, or contributes to long-term metabolic stress and neurodegenerative risk.

### MIM accurately classifies mTBI and tracks symptom burden

The multivariate interaction measure (MIM) robustly discriminated individuals with chronic mTBI from controls across multiple modeling approaches. A single-threshold decision rule (MIM > 0.174) achieved an AUC of 0.80, indicating strong sensitivity and specificity. More complex models, including penalized logistic regression and XGBoost, yielded similar performance and consistently identified elevated MIM as the most important predictor. Adding demographic variables such as age and sex provided little improvement, suggesting that MIM captures an injury-related signal independent of demographic factors. Thus, a single EEG-derived connectivity measure can accurately identify chronic mTBI, highlighting its promise as a simple and reliable screening tool.

These results are consistent with broader neuroimaging work showing that connectivity-based measures can classify TBI with high accuracy across modalities, including MEG, DTI, and resting-state fMRI.^45,48,49^ Unlike these approaches, which require high-cost imaging or high-density MEG/EEG, MIM maintained strong performance even with lower-density electrode arrays, underscoring its translational potential for clinical and field settings. Importantly, MIM also predicted individual symptom severity on the Neurobehavioral Symptom Inventory, with greater global connectivity associated with higher symptom burden. This relationship may reflect inefficient signaling or compensatory reorganization contributing to persistent cognitive and sensory difficulties. Longitudinal studies are needed to determine whether MIM changes track recovery or treatment response.

### Limitations

This study has several limitations. The modest sample size (18 mTBI, 26 controls) limits statistical power and generalizability, especially for machine learning models that typically require larger datasets. The absence of an independent validation cohort also constrains external validity, although we used rigorous analytic methods including source reconstruction, non-parametric permutation testing, and cross-validation. The cross-sectional design prevents causal conclusions about whether hyperconnectivity is compensatory, maladaptive, or static. The long post-injury duration (∼8 years) suggests these changes likely represent persistent rather than transient network alterations. We did not control for injury-related variables such as number, severity, or mechanism of injury, which may have introduced sample heterogeneity. Although detection was demonstrated with clinically relevant low-density EEG, spatial resolution may be lower than high-density systems. The consistent alterations across frequency bands and electrode configurations suggest, however, that diffuse hyperconnectivity may be a robust feature of chronic mTBI. Multimodal, longitudinal research is needed to validate and extend these findings.

### Future directions towards scalable, non-invasive biomarkers of chronic mTBI

Compared to fMRI and MEG, EEG offers greater accessibility, cost-efficiency, and scalability, making it well-suited for field-based and longitudinal applications. Our findings suggest that functional EEG metrics, particularly MIM, may serve as practical indicators of persistent network changes following mTBI, even years after injury. This durability highlights the potential of EEG not only for identifying chronic alterations but also for tracking long-term outcomes and guiding rehabilitation strategies. While we used a 128-channel system to enhance spatial resolution, the consistent effects observed with reduced montages suggest that similar findings may be achievable with portable or lower-density systems, improving feasibility in clinical and real-world settings.

Future studies should replicate these findings in larger, demographically diverse cohorts and assess whether MIM-derived features predict recovery trajectories or clinical symptoms continuously over time. Longitudinal research is essential to determine whether hyperconnectivity resolves, persists, or evolves across stages of injury and whether it correlates with cognitive or functional outcomes. Mechanistic studies integrating EEG with fMRI, PET, or diffusion MRI will be critical to determine whether elevated connectivity reflects compensatory plasticity, inefficient signaling, or ongoing neuroinflammation. If validated, EEG-based functional connectivity, measured by MIM, may offer a sensitive and scalable biomarker for detecting chronic brain changes and monitoring symptom burden, while also identifying patients who may benefit from therapeutic intervention.

## CONCLUSION

Overall, our findings demonstrate that functional connectivity, quantified by MIM, is highly sensitive to chronic mTBI. This hyperconnectivity effectively distinguishes individuals with mTBI from healthy controls and robustly predicts symptom severity across multiple machine learning approaches. Resting-state EEG combined with MIM offers an accessible, accurate tool for detecting network-level disruptions in patients with chronic mTBI. With further validation, RS-EEG could become integral to clinical classification frameworks for brain injury, such as the CBI-M model,^50^ significantly enhancing characterization and management of chronic brain injury.

## Acknowledgements

The authors would like to thank Beth Miller, Catherine Diethelm, and Logan Crawford for assistance with participant recruitment, scheduling, and performing EEG recordings for this study. The authors would also like to acknowledge that the Cartool software (cartoolcommunity.unige.ch) used in our analyses was programmed by Denis Brunet, from the Functional Brain Mapping Laboratory (FBMLab), Geneva, Switzerland, and is supported by the Center for Biomedical Imaging (CIBM) of Geneva and Lausanne.

## Funding Sources

This work was supported by the Department of Defense [W81XWH-17-2-0055, W911NF2120078, HT9425-24-1-0563, HT9425-25-1-0529]; the National Institutes of Health [NEI R01EY036252, NEI P30EY008126 (VVRC), NEI T32EY007135 (AC)]; the Marlene and Spencer Hays Directorship; the Potocsnak Discovery Grant in Regenerative Medicine; the Ret. Maj. General Stephen L. Jones, MD Fund; and the Research to Prevent Blindness, Inc. Unrestricted Funds (VEI).

## Author Contributions

WJM performed formal analysis and wrote the original draft. ANC assisted with formal analysis, supervised WJM, and reviewed and edited the writing. APK contributed to the study design, provided methodology and validation, and reviewed and edited the manuscript. TSR conceptualized the study, administered the project, provided supervision, and reviewed and edited the manuscript.

## Conflict of Interest

Authors have no competing interest to disclose.

## Transparency, Rigor, and Reproducibility Summary

The study was pre-registered with the Federal Interagency TBI registry (FITBIR; (https://fitbir.nih.gov/). The analysis plan was not formally registered, but was pre-specified in the Department of Defense grant proposal that funded this study. The plan was the responsibility of the team biostatistician. A sample size of 200 subjects per group was planned based on availability of subjects within the duration of funding. Unfortunately, due to COVID-19, total recruitment was lower than planned. Further, due to changes in personnel, EEG was not collected on the total cohort for the larger study. Thus, these data are from a subset of our cohort for the larger study: 34 control, and 28 mTBI participants. Data were collected from participants between 2017-2023, except for 2020 and 2021 due to COVID-19 pandemic restrictions. Data were collected from each participant during a single visit. Data was not analyzed until collection was completed. All data was collected from the same equipment -128-channel hydrocel net (Electrical Geodesics, Inc., Eugene, OR), and in the same location. We used MatLab software for acquisition and analysis. We relied on self-report of TBI with validation from the clinical report whenever possible. Statistical analysis performed on the EEG data was chosen based on the expertise of Dr. Key who has over 100 publications in this field. We used False Discovery Rate (FDR) correction (Benjamini & Hochberg, 1995) to correct for multiple comparisons throughout the manuscript. Planned/ongoing external validation studies have been preregistered at FITBIR. Data from this study is available in FITBIR. Analytic code used to conduct the analyses in this study is available upon reasonable request.

---

**Supplementary Figure 1.**
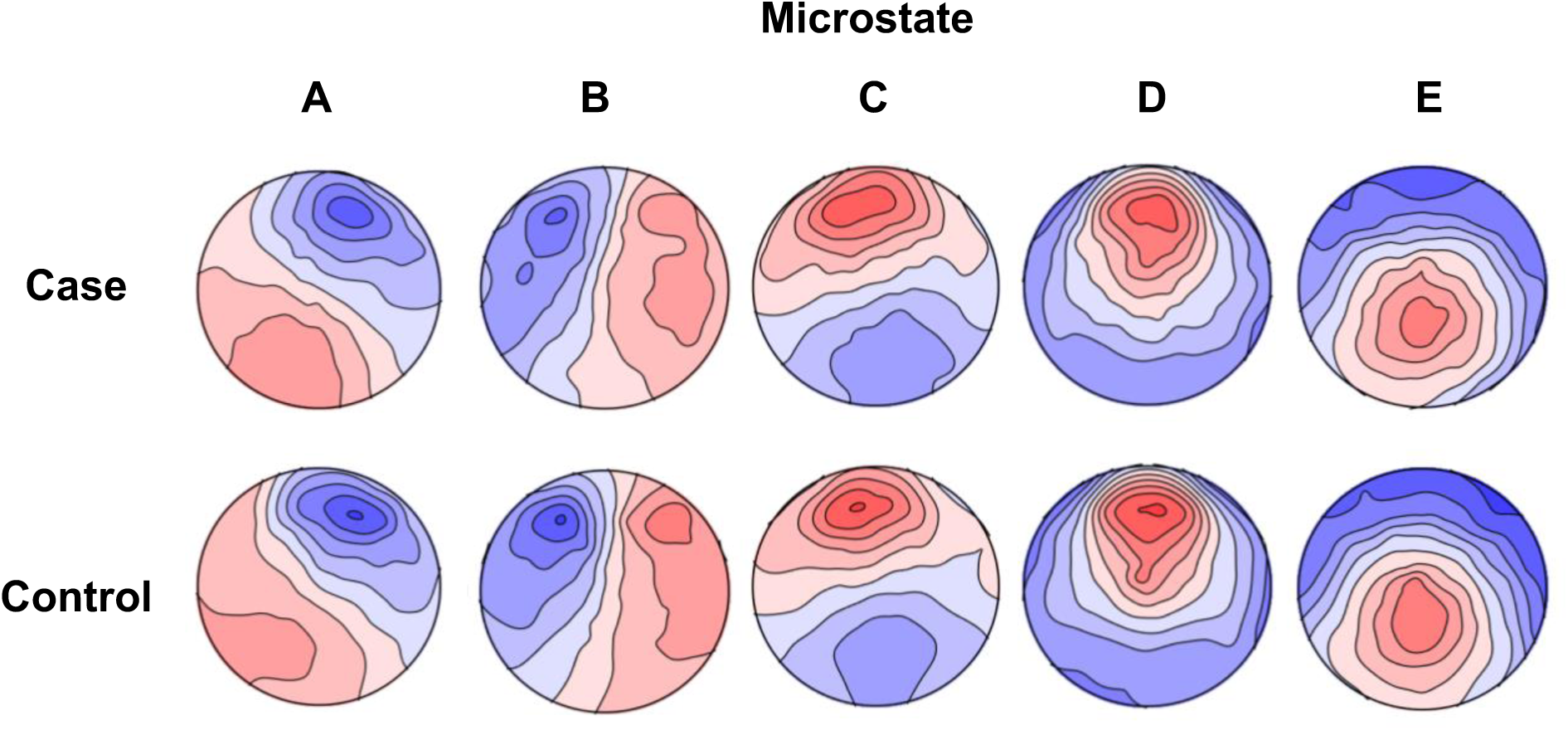
Mean Microstate Topographical Plots by Group. Topographical representations of the mean EEG microstates (A–E) for the case and control groups. Each scalp map shows the average spatial distribution of electrical potentials associated with the corresponding microstate.

**Supplementary Figure 2.**
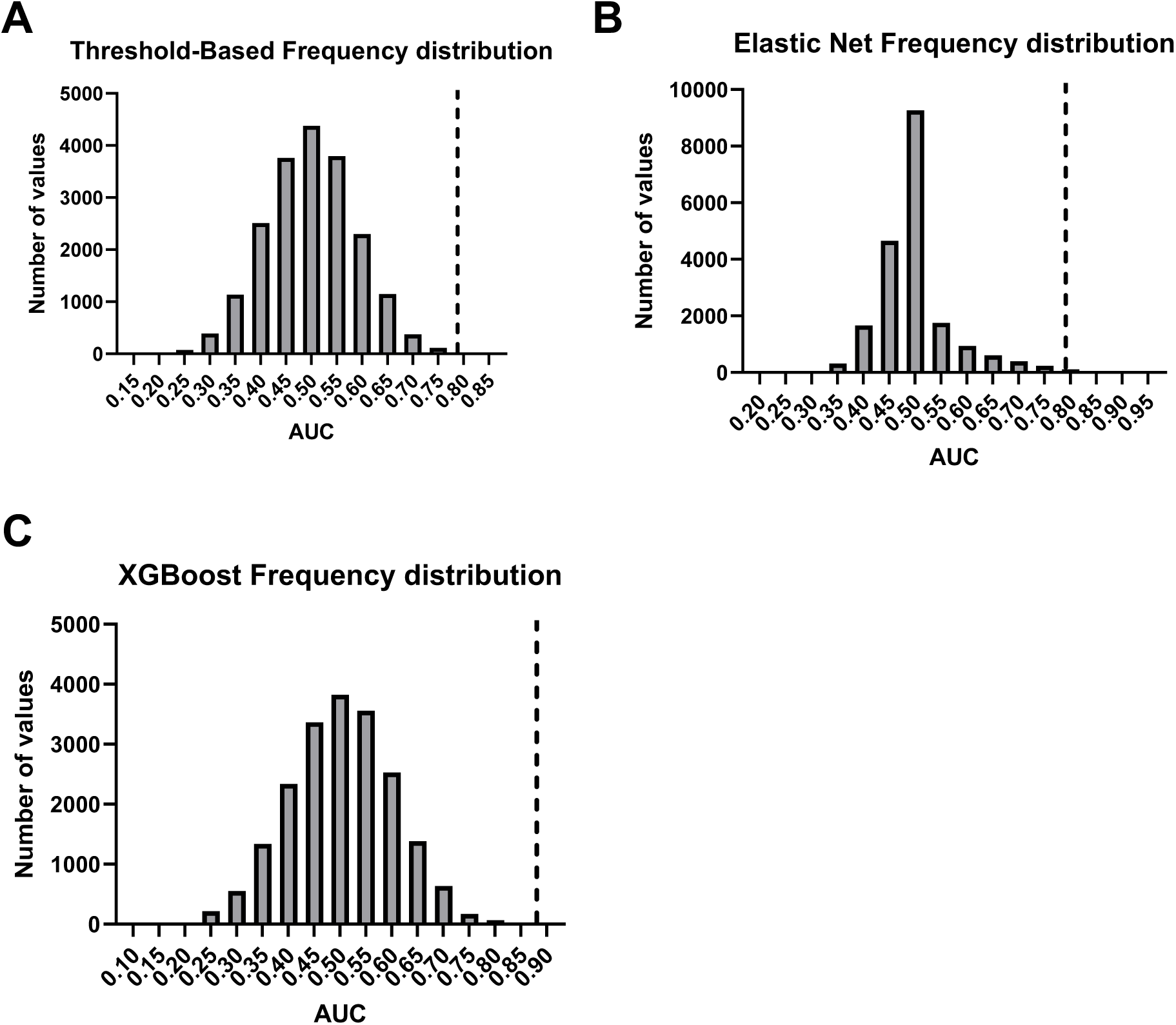
Permutation distributions of AUC (Area Under the Curve) values for each model Histograms display the frequency of AUC values under the null hypothesis for all three models (A-C). The dashed vertical line indicates the observed AUC for the true model in each case. In all comparisons, the observed AUC had statistically significant model performance above chance (p < 0.05)

**Supplementary Figure 3.**
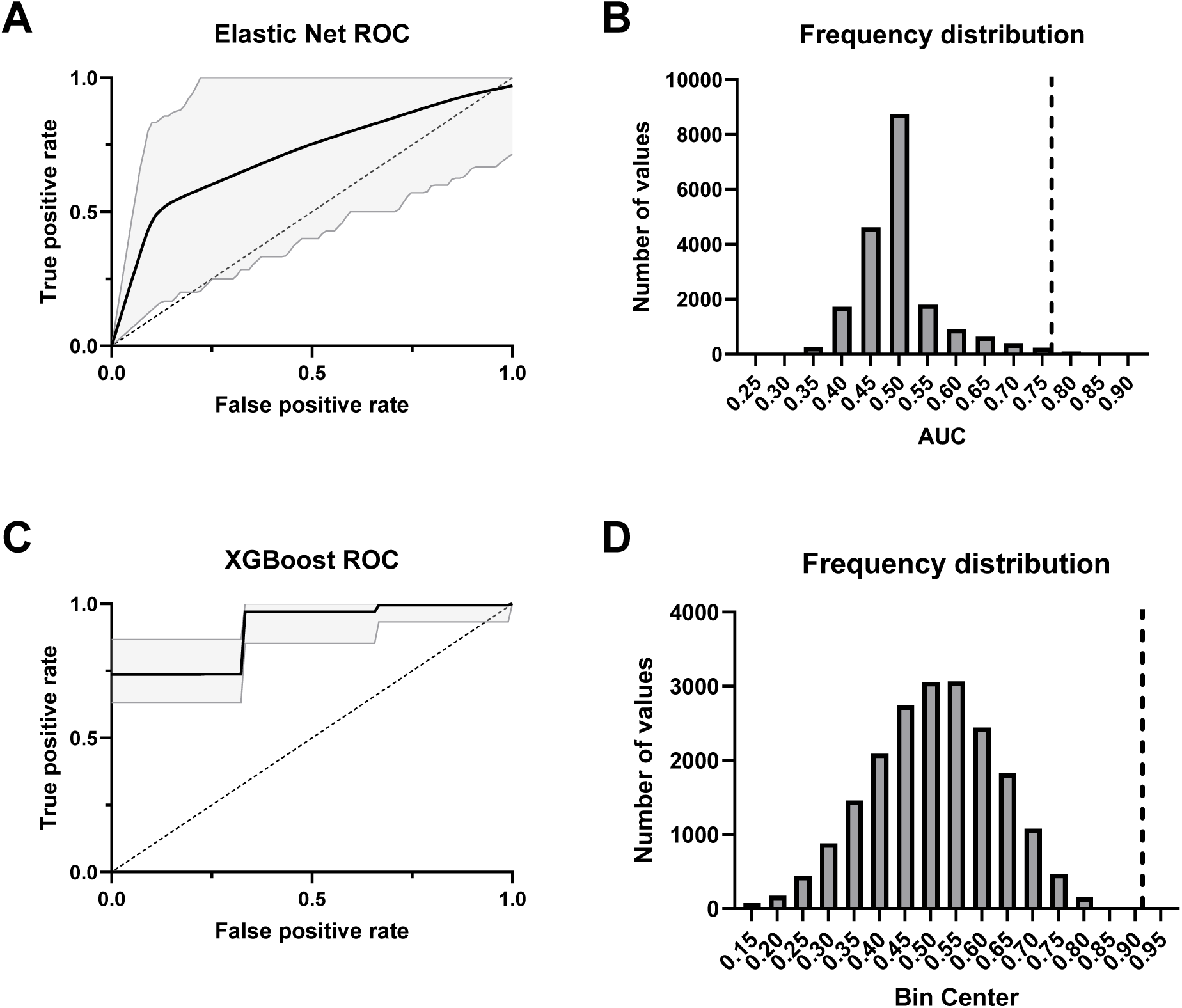
Performance of group classifier models including Age and Sex Covariates A: ROC curve for an Elastic Net logistic regression classifier trained on MIM, age, and sex. The black line indicates the mean ROC curve across 20,000 bootstraps, with 95% confidence interval shaded in gray. B: Histogram displaying the frequency of AUC values under the null hypothesis for the elastic net model. The dashed vertical line indicates the observed AUC for the true model in each case. The observed AUC had statistically significant model performance above chance (p < 0.05). C: ROC curve for an XGBoost classifier trained on MIM, age, and sex. The black line indicates the mean ROC curve across 20,000 bootstraps, with 95% confidence interval shaded in gray. D: Histogram displaying the frequency of AUC values under the null hypothesis for the XGBoost model. The dashed vertical line indicates the observed AUC for the true model in each case. The observed AUC had statistically significant model performance above chance (p < 0.05).

**Supplementary Figure 4.**
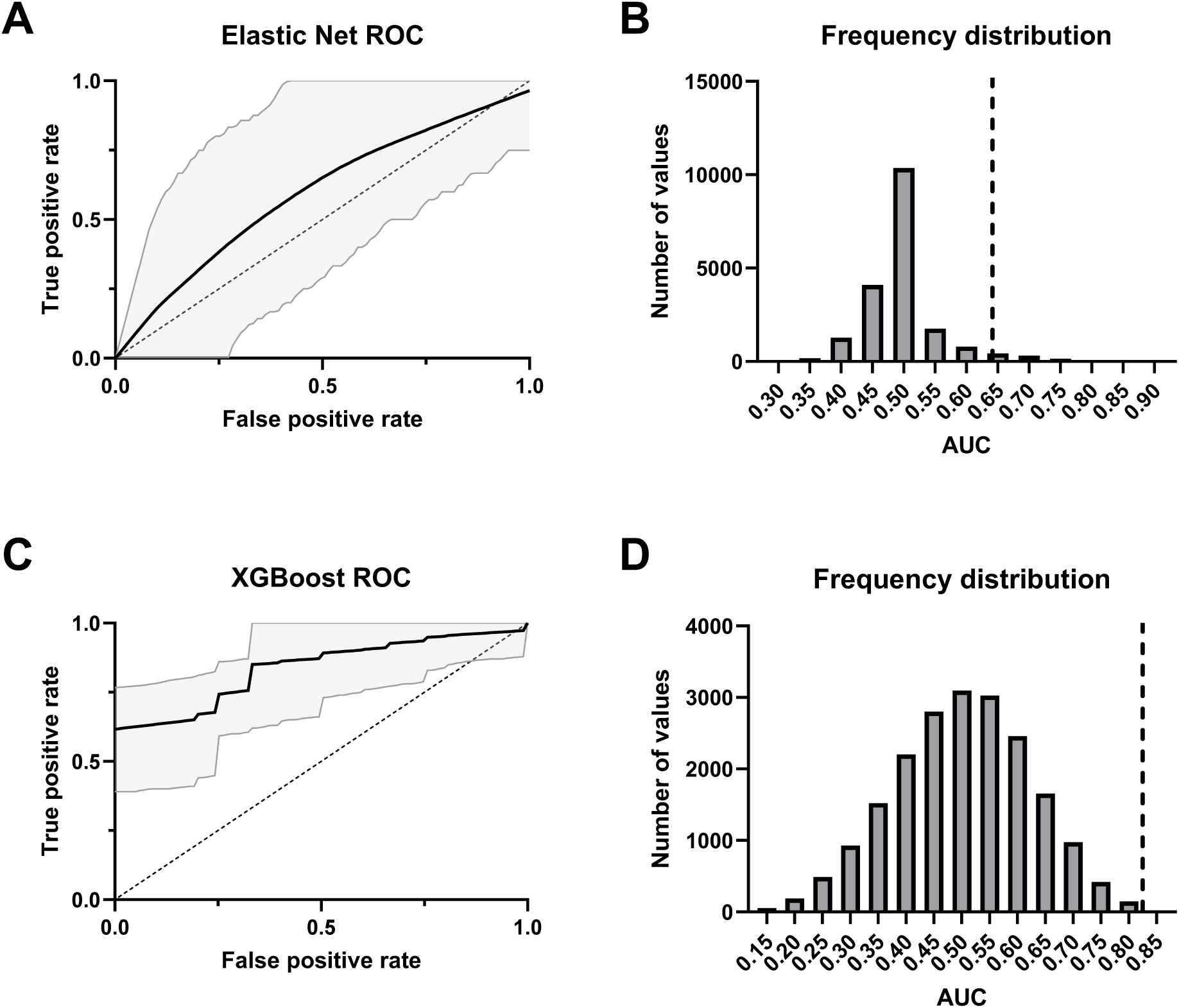
Performance of demographic group classifier models (Age and Sex Predictors only) A: ROC curve for an Elastic Net logistic regression classifier trained on age and sex. The black line indicates the mean ROC curve across 20,000 bootstraps, with 95% confidence interval shaded in gray. B: Histogram displaying the frequency of AUC values under the null hypothesis for the Elastic Net model. The dashed vertical line indicates the observed AUC for the true model in each case. The observed AUC had statistically significant model performance above chance (p < 0.05). C: ROC curve for an XGBoost classifier trained on age and sex. The black line indicates the mean ROC curve across 20,000 bootstraps, with 95% confidence interval shaded in gray. D: Histogram displaying the frequency of AUC values under the null hypothesis for the XGBoost model. The dashed vertical line indicates the observed AUC for the true model in each case. The observed AUC had statistically significant model performance above chance (p < 0.05).

**Supplementary Figure 5.**
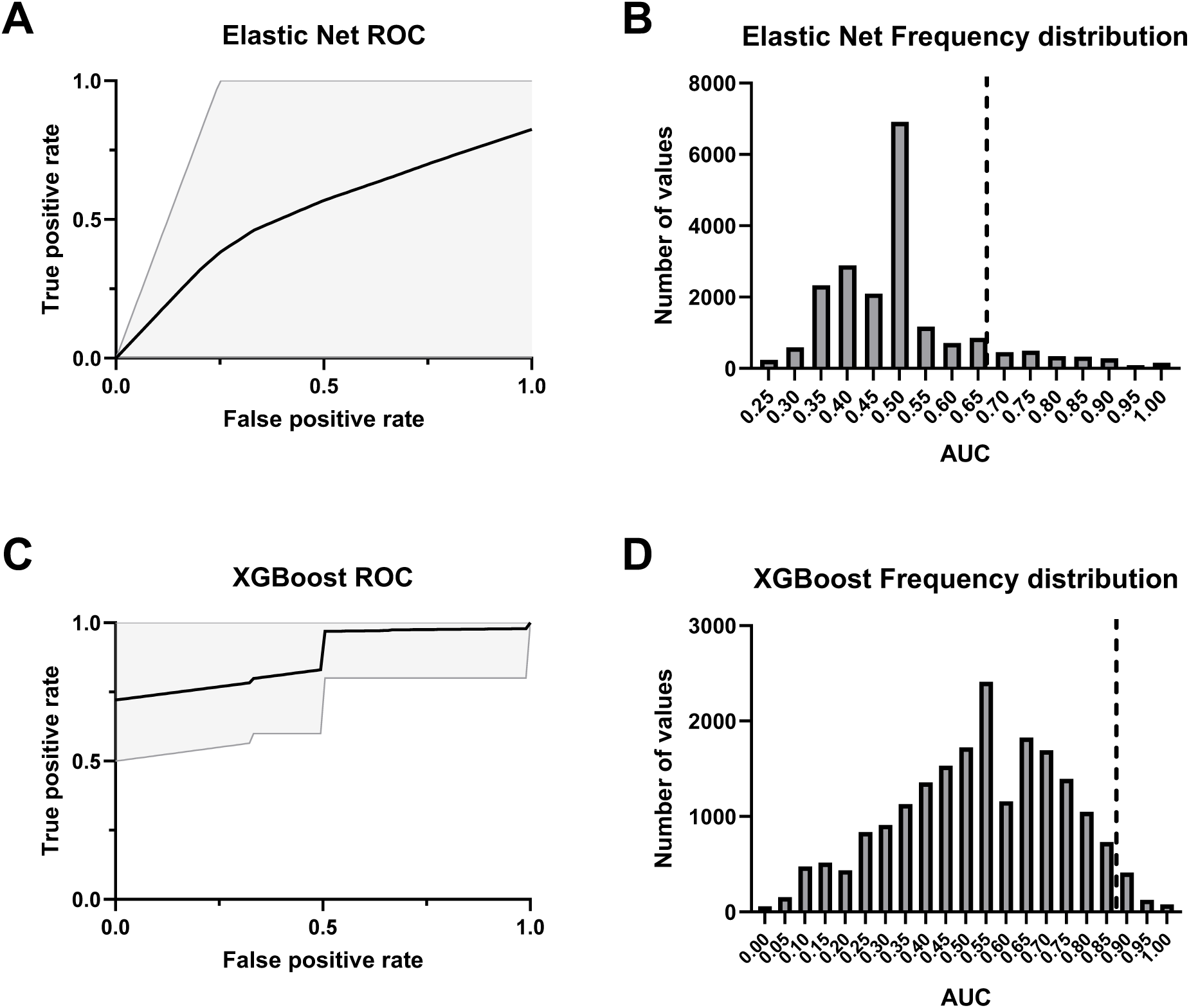
Performance of symptom classifier models including Age and Sex Covariates A: ROC curve for an Elastic Net logistic regression classifier trained on MIM, age, and sex. The black line indicates the mean ROC curve across 20,000 bootstraps, with 95% confidence interval shaded in gray. B: Histogram displaying the frequency of AUC values under the null hypothesis for the elastic net model. The dashed vertical line indicates the observed AUC for the true model in each case. The observed model did not perform significantly above chance (p = 0.125). C: ROC curve for an XGBoost classifier trained on MIM, age, and sex. The black line indicates the mean ROC curve across 20,000 bootstraps, with 95% confidence interval shaded in gray. D: Histogram displaying the frequency of AUC values under the null hypothesis for the XGBoost model. The dashed vertical line indicates the observed AUC for the true model in each case. The observed AUC had statistically significant model performance above chance (p < 0.05).

**Supplementary Figure 6.**
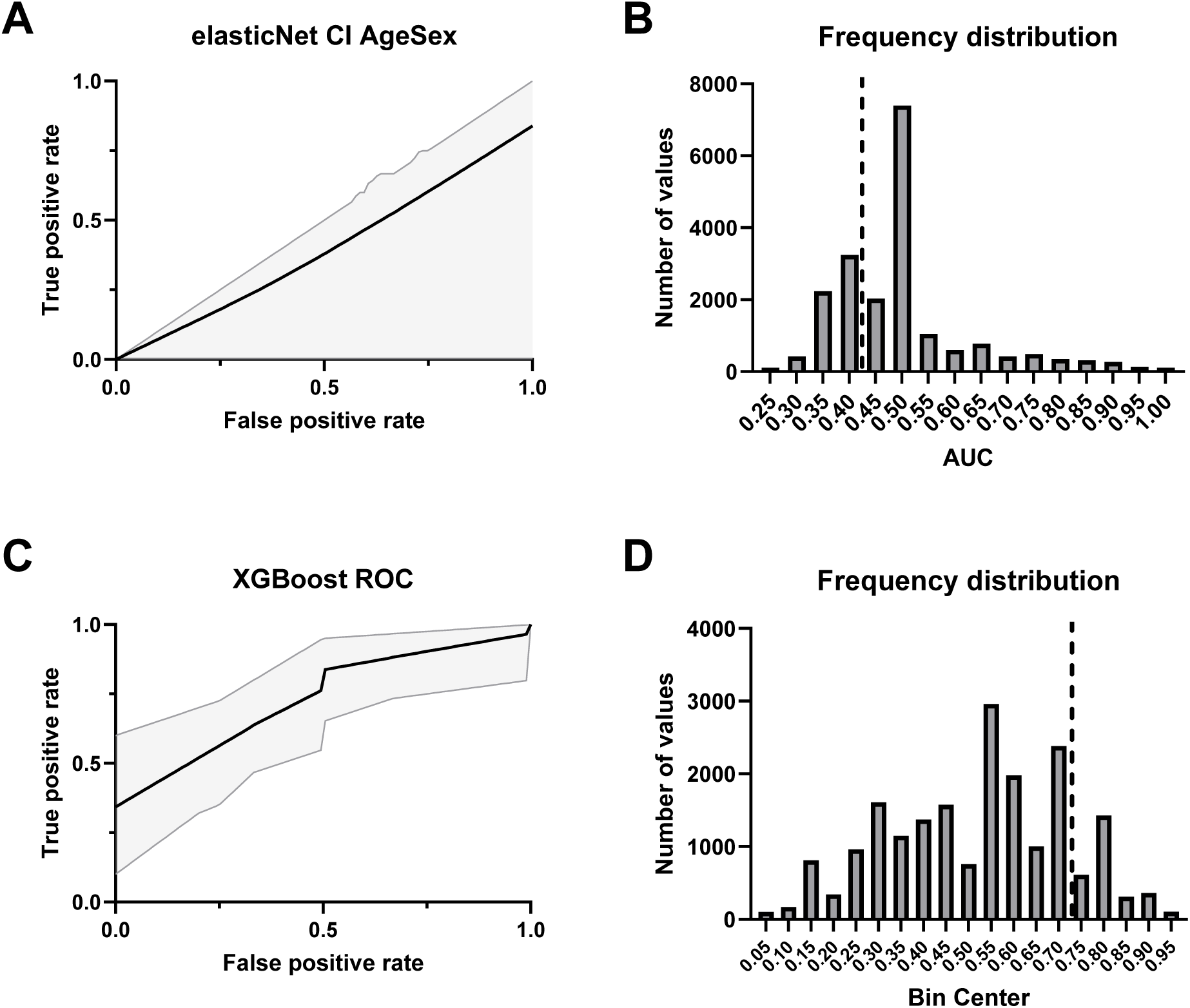
Performance of symptom classifier models trained on demographics. A: ROC curve for an Elastic Net logistic regression classifier trained on age and sex. The black line indicates the mean ROC curve across 20,000 bootstraps, with 95% confidence interval shaded in gray. B: Histogram displaying the frequency of AUC values under the null hypothesis for the Elastic Net model. The dashed vertical line indicates the observed AUC for the true model in each case. The observed model did not perform significantly above chance (p = 0.698). C: ROC curve for an XGBoost classifier trained on age and sex. The black line indicates the mean ROC curve across 20,000 bootstraps, with 95% confidence interval shaded in gray. D: Histogram displaying the frequency of AUC values under the null hypothesis for the XGBoost model. The dashed vertical line indicates the observed AUC for the true model in each case. The observed model did not perform significantly above chance (p = 0.147)

**Supplementary Table 1.** Participant Demographics. mTBI = mild traumatic brain injury.

| Variable | Control (n = 26) | mTBI (n = 18) |
| --- | --- | --- |
| Age, mean (SD) | 39.6 (11.5) | 32.0 (11.4) |
| Sex, % Female | 73% | 50% |
| **# of mTBIs, n (%)** |  |  |
| 1 | — | 9 (50%) |
| 2 | — | 1 (6%) |
| 3 | — | 6 (33%) |
| 4 | — | 2 (11%) |
| Did not report | — | 0 (0%) |
| **Years Since Last mTBI, n (%)** |  |  |
| < 1 | — | 1 (6%) |
| 1–2.9 | — | 6 (33%) |
| 3–6.9 | — | 2 (11%) |
| 7–9.9 | — | 2 (11%) |
| > 10 | — | 6 (33%) |
| Did not report | — | 1 (6%) |

## Notes

### Competing Interest Statement

The authors have declared no competing interest.

### Author Declarations

The Institutional Review Board of Vanderbilt University Medical Center (IRB #171061) gave ethical approval for this work

